# Landscape of parental postzygotic mutations in >11,000 rare disease trios

**DOI:** 10.1101/2025.10.17.25337713

**Authors:** O. Isaac Garcia-Salinas, Rashesh Sanghvi, John A. Sayer, Maria Torra I Benach, My H. Pham, Aylwyn Scally, Hilary C. Martin, Raheleh Rahbari

## Abstract

Postzygotic mutations (PZMs) arising post-fertilisation, prior to primordial germ cell specification, may be subsequently inherited by both somatic and germ cells, causing somatic mosaicism in the parent as well as being passed to offspring. These early embryonic mutations often go undetected in clinical sequencing due to their low variant allele fraction (VAF) and are typically excluded by standard variant filtering pipelines. To overcome this limitation, we developed a bioinformatic approach to detect PZMs using standard-depth (∼30X) whole genome sequencing data from 12,015 parent-offspring trios in the Genomics England 100,000 Genomes Project. We identified 1,015 high-confidence autosomal early PZMs. These mutations showed no parental sex-bias and exhibited a monomodal VAF distribution centred around 5% in blood. PZMs displayed a mutational spectrum distinct from *de novo* mutations, although both appeared to be driven by mutational signatures SBS1 and SBS5. Notably, we identified a subset of PZMs likely contributing to the clinical phenotype in affected children. This work demonstrates that postzygotic mosaicism represents a rare but clinically relevant source of diagnostic variation in rare disease and can be detected from standard sequencing data. Incorporating PZM detection into clinical workflow could improve diagnostic yield and provide more accurate recurrence risk estimates for affected families.

## Main Text

Most heritable genetic mutations arise in the germ cells during gametogenesis across the lifetime of an individual and are known as *de novo* mutations (DNMs)^1,2^. However, mutations can also occur earlier in development, including as early as the first zygotic cell division. These are termed postzygotic mutations (PZMs)^1,3,4^. Depending on developmental timing, PZMs can be classified into two categories. Early PZMs arise before somatic and germline fates are specified (approximately two weeks post-fertilization and within the first 10–15 cell divisions)^5,6^ and therefore generate mosaicism in both compartments. Germline-confined PZMs occur later in germline progenitors and are restricted to the germline lineage. Importantly, the level of mosaicism in fully developed tissues reflects the timing of the mutation, with earlier events present at higher cell fractions and more likely to recur in the carrier’s future offspring ^7,8^ (**Figure 1A–B**).

**Figure 1.**
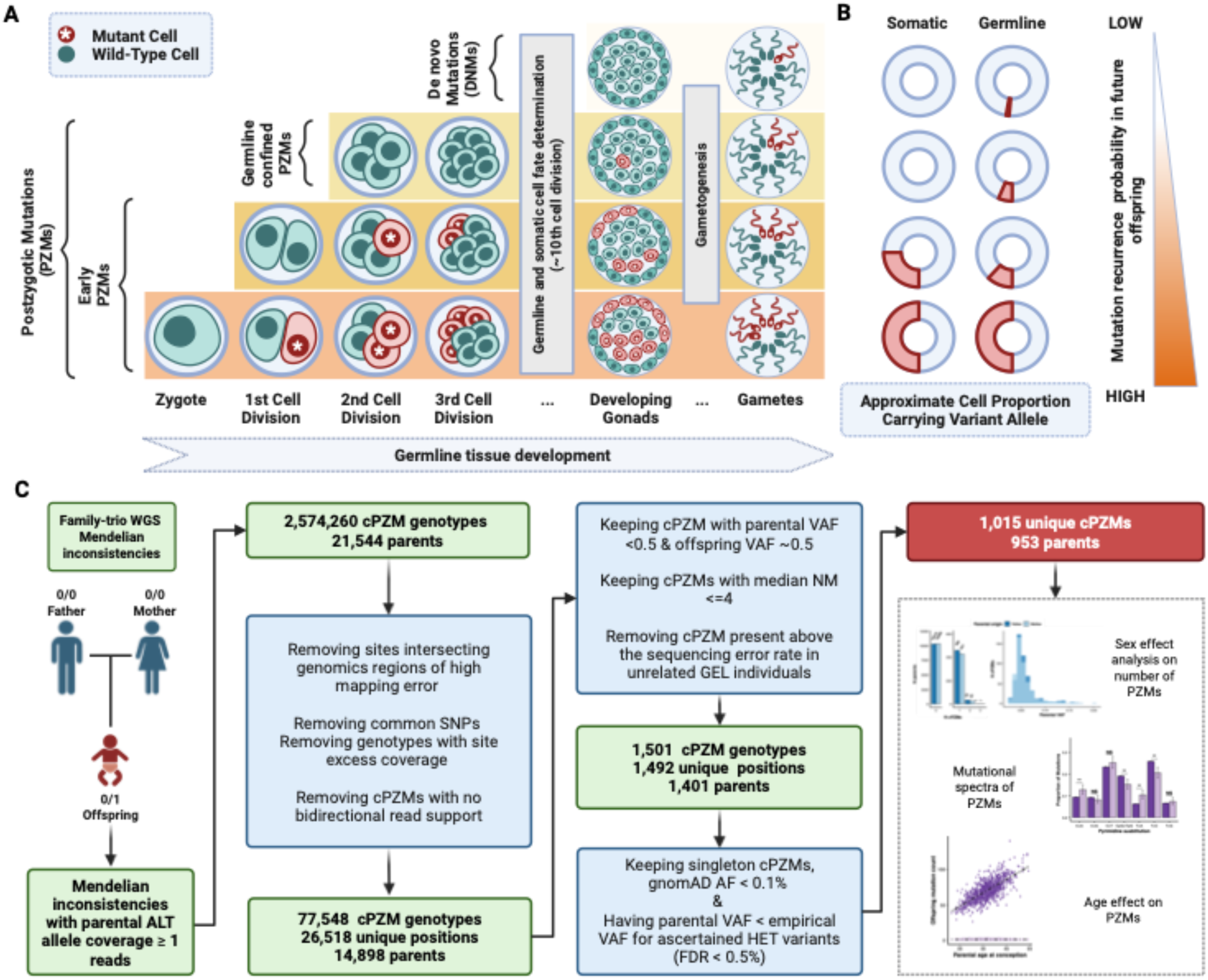
Origins of germline mutations and bioinformatic ascertainment of parental postzygotic mutations. **A.** Classification of germline mutations according to mutation timing. Postzygotic mutations (PZMs, rows 1–3, bottom to top) can affect both somatic and germline lineages if they occur before segregation of somatic and germline fates (i.e., early PZMs), or be restricted to the germline tissue if they arise in germline progenitors (i.e., germline-confined PZMs). For both categories, the earlier a mutation occurs, the larger the fraction of future gametes that will carry it. In contrast, *de novo* mutations (DNMs, top row) arise later in life during gametogenesis and affect only a subset of germline progenitors or single gametes. Gametes are represented with sperm cells, however, germline mutations can arise in both the male and female germline. **B.** Approximate proportions of somatic and germline cells carrying each type of mutation, with the right-hand scale indicating the likelihood of transmission to offspring. **C.** Overview of this study: analysis of trio WGS data for Mendelian inconsistencies supported by ≥1 ALT read, yielding 1,015 candidate early PZMs (cPZMs) in parents.

Although PZMs are a recognized source of sporadic genetic disease^4,8,9^, they are frequently overlooked by standard germline variant-calling pipelines because their variant allele fraction (VAF) is lower than expected for a heterozygous germline variant in the sequenced tissue^10^. This issue is particularly relevant in clinical sequencing of family trios with rare diseases, typically comprising an affected child and two unaffected parents. In such cases, a pathogenic parental PZM may be overlooked since the variant will have too high a VAF to be called as a DNM or, less commonly, because the mosaic parent is affected but the variant has too low a VAF to be called as heterozygous in them^7,8^.

A possible approach to identify parental PZMs in parent-offspring trio sequencing studies is to analyse the parental VAF of candidate DNMs. When a candidate DNM has non-zero coverage for the alternative allele in a parent, it suggests that the mutation may have arisen postzygotically in the parent and was then transmitted to the offspring^4,11^. Identifying parental PZMs through this approach presents several challenges. First, because PZMs are present in smaller subsets of cells, they may not be reliably detected by standard variant-calling methods in standard-depth whole genome sequencing (WGS) due to the high sequencing error rate. Low VAF variants are often due to artefacts, particularly in regions with poor coverage or plagued by sequencing and mapping errors^11,12^. Annotations incorporating statistical comparison of features such as read depth, allele balance, and quality scores can be exploited to distinguish between real mosaic variants and sequencing errors^13^.

While many studies have centred on identifying PZMs through trio WES, most of these have typically focused on ascertaining pathogenic coding PZMs in rare disease patients^8,14–16^. This limited the ability of these studies to characterise PZMs at scale due to their sample size and focus on the protein-coding genome in which the yield of PZMs is low. Only four studies to date have examined PZMs using WGS data^1,3,4,17^. However, these efforts have been constrained by limited sample sizes, typically ranging from 6 to ∼1000 offspring^1,3,17^. The largest catalogue of PZMs to date came from the study by Sasani et al. (2019) who sequenced 70 three-generation families, detecting 1,195 PZMs using two complementary strategies: one biased towards PZMs detectable only in germline tissues and occurring early enough to be shared between siblings (n = 720), and another detecting early PZM events likely occurring in developmental progenitors of both somatic and germline tissues through phasing and co-segregation analyses across three generations (n = 475)^17^. They found that the mutation spectrum of early and germline-confined PZMs differ from conventional DNMs, with early PZMs showing a depletion in T>A changes relative to DNMs, and germline-confined PZMs showing a depletion in T>C and enrichment in C>A and CpG>TpG changes. While informative, these methods are difficult to scale and are not applicable to standard two-generation trios.

To address this, we developed a novel approach that relies on filtering candidate DNMs within two-generation trios, focusing on those in which the alternate allele is detectable in parental reads while minimising artefacts, enabling the detection of early PZMs, i.e., mutations arising before germline and somatic lineages separate, in large cohorts. The Genomics England (GEL) 100,000 Genomes Project (100kGP) is an initiative aiming to identify the genetic causes of rare diseases through WGS in patients from the United Kingdom^18^. As part of this endeavour, the 100kGP identified DNMs across 12,015 trios, generating one of the largest germline mutation catalogues to date. Importantly, the analysis of germline variation in GEL trios did not include any methods for the detection of postzygotic mutation events in trio parents, which led to the misclassification of these variants as either DNMs in the child (presumed to occur only in the parental germline) or technical artefacts. Applying our pipeline to the 100kGP trios, we identified 1,015 candidate early PZMs (**Figure 1C**), characterised their mutational spectrum and genomic distribution, and identified putatively pathogenic variants.

We accessed a previously identified set of 5,720,018 autosomal single nucleotide variants (SNVs) exhibiting Mendelian inconsistencies across an initial set of 11,996 trios (involving 21,644 unique parents) recruited through the rare disease arm of the 100k Genomes Project (100kGP)^19^ (**Supplementary Methods**). Mendelian inconsistencies were defined as sites where the offspring was called as heterozygous for the alternative allele, while both parents were called as homozygous for the reference allele. These candidate DNMs were identified on a per-trio basis using the variant caller Platypus, combining local realignment, haplotype assembly, and Bayesian statistical modeling^20^.

Among these, 2,627,735 candidate autosomal SNV DNMs had alternate allele read support in a single parent, suggesting possible parental postzygotic mosaicism. We refer to these variants as candidate parental postzygotic mutations (cPZMs). Since multiple offspring from the same parent could appear in different trios, we de-duplicated such genotypes, yielding an initial set of 2,574,260 cPZMs (mean = 118.93 cPZMs per parent). This set was then subjected to a custom filtering pipeline to distinguish true PZMs from sequencing artefacts and common polymorphisms (**Supplementary Methods**; **Supplementary Figure 1**), obtaining 1,015 high-confidence cPZMs, corresponding to 0.039% of the initial candidates (**Supplementary Figure 2**).

After accounting for individuals removed due to missing metadata (n = 123, **Supplementary Methods**), we identified an average of 0.046 PZMs per parent. This is a markedly lower rate than previously reported whole-genome estimates such as Rahbari et al., (2016)^1^ who reported an average of 4 PZMs per parent, while Sasani et al. (2019)^17^ reported a mean of 6.40 per parent (**Supplementary Methods**). Our estimate is likely more conservative due to key technical limitations as we relied on standard sequencing depth and lacked large scale multigenerational or sibling data, both of which substantially enhance sensitivity to detect variants with lower somatic VAF or those with a VAF closer to 0.5, respectively. Despite this limitation, our catalogue spans the largest number of individuals and the largest number of early PZMs detected in a single study, allowing us to characterise PZMs at scale (**Figure 2**).

**Figure 2.**
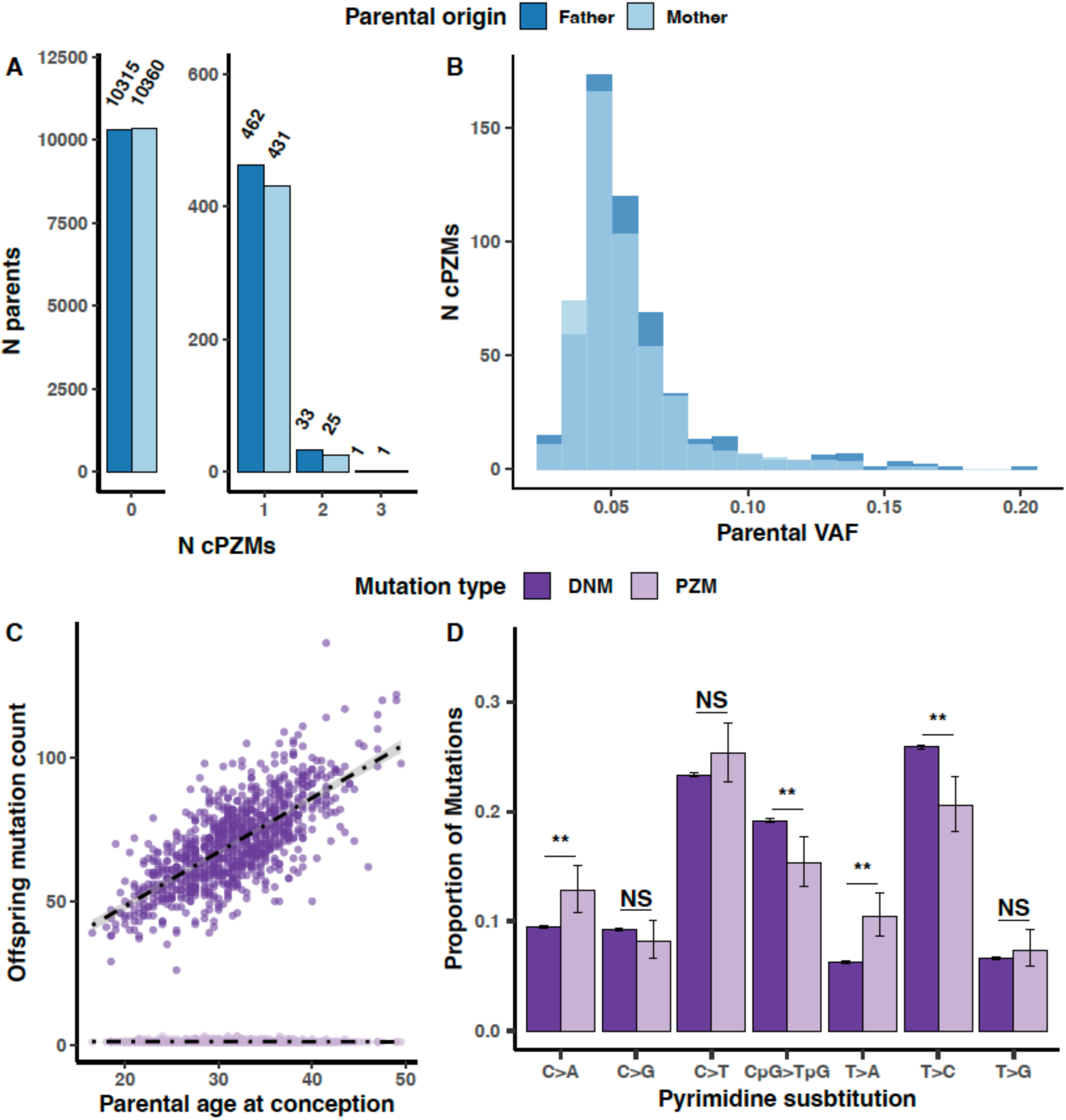
Characteristics of parental postzygotic mutations. **A.** Distribution of PZM burden (number per individual) across the parental cohort (n = 21,650). **B.** Distribution of PZM variant allele fraction (VAF). No significant maternal vs paternal differences in burden or VAF (Wilcoxon rank-sum p = 0.19 and 0.09, respectively). **C.** Effect of mean parental age at conception on offspring PZMs (light purple) and DNMs (dark purple). No significant age effect for PZMs (generalised linear regression p = 0.98). **D.** Mutation spectra comparison between PZMs and DNMs. PZMs were enriched for T>A and C>A mutations (two-sample proportion test p = 2.1 × 10⁻³; 4.7 × 10⁻⁷) and depleted for CpG>TpG and T>C (FDR-adjusted p = 1.3 × 10⁻²; 8.9 × 10⁻⁴) compared to DNMs.

We observed no significant difference in the number of PZMs identified between parental sexes (Wilcoxon rank-sum test p-value = 0.19, **Figure 2A**), with 52.3% of the total mutation events identified in fathers. This suggests that in humans, the opportunity to acquire early PZMs is similar across sexes. In contrast, model organisms such as mice exhibit a paternal bias in this type of mutations, likely due to earlier primordial germ cell (PGC) specification in males^21^. Thus, our results suggest that these sex-specific developmental mechanisms observed in mice do not operate in the same way in humans.

Across all cPZMs, we observed a monomodal distribution of VAFs centred on 0.05 (±2.77 × 10^-3^, 95% confidence interval [CI]), which did not differ significantly between parental sexes (Wilcoxon rank-sum test p-value = 0.09, **Figure 2B**). Under a simple model of synchronous cell divisions, stable diploidy, no selection or cell loss, and uniform contribution of daughter lineages, most mutations (n = 662) would be inferred to have arisen after the third embryonic division (**Supplementary Table 1**). However, this interpretation should be treated with caution.

If some early lineages were lost, contributed disproportionately, or were excluded from the embryoblast, the relationship between VAF and cell-division timing would be distorted. The observed distribution likely reflects a combination of biological processes and technical constraints, including our deliberate exclusion of high-VAF variants (to avoid including germline heterozygotes) and limited sensitivity for low-VAF variants at standard sequencing depth. As such, our dataset is likely underpowered to detect both the earliest PZMs (1st-2nd cell division) and those arising close to PGC specification (∼10-15th division), after which PZMs remain confined exclusively to the germline tissues.

The parental age effect on DNM rate is well-established, and has been proposed to result from the interplay between DNA damage and repair throughout ageing^22^. However, since PZMs arise during early development, we do not expect them to be associated with parental age. To test this, we aggregated PZM counts across both parents for each offspring and modelled the total as a function of maternal and paternal age at conception, adjusting for technical covariates **(Supplementary Methods; Figure 2C)**. As expected, neither maternal nor paternal age showed a significant effect on PZM counts (p-value = 0.5 and 0.42, respectively). In contrast, DNM counts were significantly and positively associated with both maternal age (effect = 7.2 x 10^-3^, p-value = 6.37 x 10^-14^) and paternal age (effect 1.8 x 10^-2^, p-value < 2 x 10^-16^). These results support the specificity of our PZM calls and suggest that they are not confounded by misclassified DNMs or by parental-age-related somatic variants.

To investigate whether postzygotic mutations (PZMs) exhibit non-random genomic distributions, potentially reflecting the influence of genomic features or developmental processes, we partitioned the genome into 2 Mb bins and tested whether any showed a significant excess of PZMs relative to a uniform null model (**Supplementary Methods**). We identified 49 bins with nominally significant enrichment (one-tailed empirical p ≤ 0.05; **Supplementary Figure 3**), although none remained significant after multiple testing correction (FDR > 5%). With this caveat, we asked whether the nominally enriched bins were associated with GC content or replication timing (**Supplementary Methods**). Using binomial regression while adjusting for callable size, we found that GC content was negatively associated with PZM enrichment (odds ratio [OR] = 0.87, p = 0.016), whereas replication timing showed no significant effect (p = 0.51). Applying the same methods to bins enriched for DNMs, we observed a positive association with GC content (OR = 1.08, p = 2.15 × 10^-4^) and a negative association with replication timing (OR = 0.60, p = 9.58 × 10^-7^). While we are cautious on interpreting the results for PZMs due to our limited power, the findings on DNMs are consistent with prior reports, lending confidence in the robustness of our approach^23,24^.

Next, we investigated the characteristics of the PZM mutational spectra by classifying variants into six substitution types, namely C>A, C>T, C>G, T>A, T>C, T>G which are based on the pyrimidine substitution caused by each single point mutation^25^. We additionally considered C>T substitutions occurring in CpG sites (CpG>TpG) to account for the increased number of transitions occurring at CpGs due to spontaneous cytosine deamination^1,26^. We first compared substitution type proportions between paternal and maternal PZMs, finding no significant differences between the two groups (two-sample proportion test, FDR-adjusted p-value > 0.05; **Supplementary** Figure 4). These results are consistent with our observed lack of sex differences in mutation counts per individual, and support the hypothesis that mutagenic processes causing postzygotic mutations in the early differentiating embryo are homogeneous across sexes.

We then compared the mutational spectra of PZMs and DNMs, identifying four substitution classes with significant differences in relative frequency (two-sample proportion test, FDR-adjusted p ≤ 0.05). Specifically, PZMs were significantly enriched for C>A and T>A mutations (PZM/DNM FC = 1.36; 1.67, adjusted p = 2.12 × 10^-3^; 4.7 × 10^-7^, respectively) relative to DNMs, and significantly depleted for CpG>TpG and T>C mutations (PZM/DNM FC = 0.797; 0.794, adjusted p = 1.3 × 10^-2^; 8.9 × 10^-4^, respectively). The enrichment of C>A and T>A, and depletion of T>C are consistent with findings reported by Sasani et al. (2019) ^17^, although with some caveats. The T>A enrichment they reported was specific to their early PZM set, while the C>A and T>C enrichments were specific to their germline-confined mosaic group. Importantly, Sasani et al. did not apply any method for detecting low-VAF mosaicism in parental blood for their germline-confined mosaic group, relying instead on standard germline variant calling and genotyping methods. It is possible that a subset of their germline confined mosaic calls are actually early PZMs that simply had very low-VAF in parental blood. This could explain why our single set of PZMs captures both the early (T>A) and later (C>A, T>C) mutational patterns reported as distinct in Sasani et al.

To identify mutational processes leading to the observed spectrum differences between PZMs and DNMs, we performed mutational signature extraction and deconvolution **(Supplementary Methods).** Briefly, this analysis involves dimensionality reduction and unsupervised clustering of mutation counts across all possible pyrimidine substitution types and sequence flanking contexts^25^, which facilitates the identification of mutational patterns that correspond to known and unknown biological and environmental mutagenic factors^25^. While applying this technique to counts of PZMs and DNMs pooled across individuals, we found that “clock-like” mutational processes, namely SBS1 and SBS5^27^, underlie both mutation types (**Supplementary Figure 5**). The effect of these processes is known to be widespread across somatic^25,28^ and germline tissues^1^. The signatures are likely to originate from unrepaired errors during normal DNA processing and replication^22^.

Although mutational processes such as SBS1 and SBS5 operate across different contexts and cell types, their relative contributions and sequence-context sensitivities can vary during development and across cell types (e.g., genetically driven germline hypermutation^29^). When we compared the proportion of mutations attributed to SBS1 and SBS5 between PZMs and DNMs, we found no significant evidence for a difference (two-sample proportion test, p-value = 0.39 and 0.47, respectively). This suggests that both mutation types are largely shaped by the similar mutational processes under normal conditions. However, the substitution-level differences we observed likely reflect subtle developmental or context-specific shifts in the activity of these processes rather than entirely distinct mutational origins.

Reflecting the sequence composition of the genome, *in silico* consequence prediction^30^ showed that 96.6% of PZMs occurred in non-exonic locations (**Supplementary Figure 6**). We assessed whether PZMs were preferentially located in specific functional genomic contexts, namely transcriptional units (i.e., introns, exons, UTR regions), putative regulatory regions (5000 bases upstream transcription start sites or downstream transcription termination sites), and intergenic regions (**Supplementary Methods**), but we could not find any greater-than-expected overlap between these PZMs and any of the tested functional regions. The VAF of PZMs was also not significantly different between consequence classes.

One of the main goals of GEL is the identification of disease-causing genetic variation in patients with rare diseases^19^. This work involves identifying likely pathogenic protein-coding variants that match a relevant mode of inheritance in a set of genes considered likely to be related to the patient’s phenotypes^18,31^, termed “green panel” genes in the PanelApp tool^18,31^.

In our PZM catalogue, 917 of the identified mutations (90.34%) were filtered out by standard *de novo* filtering pipelines because they showed some low-level parental alternative allele reads (**Supplementary Methods**). These were therefore excluded from diagnostic consideration. The remaining 98 variants had no detectable supporting allele in the parents and were originally classified as DNMs by GEL. Our results show that some of these DNMs were in fact postzygotic mutations present at low-VAF in one of the parents. Among these reclassified DNMs, we identified a missense variant in *VAMP2*, which had been reported as the diagnostic variant in a patient with specific learning disability and global developmental delay. Although this finding could in theory have had clinical relevance for the family (by informing recurrence risk), the parents of the mosaic individual were past reproductive age, so it was decided not to report it back to the family.

Extending this approach, we intersected genes carrying protein-coding PZMs with a VEP “MODERATE” or “HIGH” consequence with PanelApp green genes, and identified four additional variants in genes with established clinical relevance: *WT1, DYNC1H1, PHIP,* and *NLRP3*. For each, we reviewed the diagnosis status of the proband, examined their recorded phenotypes, and compared these with gene-disease associations documented in PanelApp and the literature. The proband carrying the *NLRP3* variant had already received a diagnosis based on another pathogenic variant, and the *PHIP* variant carrier showed only a limited match to expected phenotype for this gene. The remaining two variants, in *WT1* and *DYNC1H1*, had potential clinical relevance for previously undiagnosed patients (**Table 1**).

**Table 1.** Summary of clinically relevant parental PZMs in the GEL cohort.

| Gene | HGVS ID | Parental VAF | Patients phenotypes explained | Notes |
| --- | --- | --- | --- | --- |
| *VAMP2* | ENST00000316509.11:c.197G>C | 0.0308 | Specific learning disability, Global developmental delay | Variant already reported as a pathogenic DNM |
| *DYNC1H1* | ENST00000360184.10:c.1705C>T | 0.0526 | Bilateral tonic-clonic seizure | Same variant occurred DNM in another GEL patient and was determined to be pathogenic; clinician unavailable to compare patients' phenotypes so pathogenicity of this PZM uncertain |
| *WT1* | ENST00000452863.10:c.1455G>T | 0.0333 | Hydroureter, Vesicoureteral reflux, Proteinuria | Variant considered clinically reportable |

The missense variant in *DYNC1H1* affects the canonical transcript (ENST00000360184.10, exon 8/78), is not reported in gnomAD, and was classified as likely pathogenic by ClinPred and AlphaMissense (scores 0.97; 0.99, respectively), and mildly pathogenic by CADD (score 24.8). The affected gene is associated with peripheral neuromuscular and neurodevelopmental disorders^32,33^, including epilepsy^34^. It was identified in a patient with bilateral tonic-clonic seizures, autism and macrocephaly. The same variant had also been identified as diagnostic in another unrelated GEL participant who also had seizures but otherwise non-overlapping features, including cerebral palsy and hemiplegia. Although initially classified as a DNM in that individual, its VAF was significantly lower than 0.5 (VAF = 0.31, one-tailed binomial test, p = 0.025), suggesting mosaicism and indicating that the mutation may have arisen during early embryonic development. The clinical team for our index case regarded the variant as potentially pathogenic, but attempts to establish phenotypic concordance with the second case were unsuccessful. Further characterisation and further genotype-phenotype data from additional cohorts will be required to clarify the significance of this variant and its potential role as mosaic contributor to disease.

Finally, we identified a postzygotic missense variant in *WT1*. This variant affects the gene’s canonical transcript (ENST00000452863.10, exon 10/10), is absent from gnomAD, and showed moderate to high predictions of deleteriousness by CADD (23.4), REVEL (0.31), and ClinPred (0.99). This is a gene in which mutations are known to cause Wilms tumor and gonadal and urinary tract malformations^35^. The carrier case displayed phenotypes indicative of congenital anomalies of kidney and urinary tract (CAKUT), including hydroureter, vesicoureteral reflux, and proteinuria. Although *WT1* was not included in the CAKUT-associated gene panel used for clinical diagnosis in this individual, vesicoureteral reflux and proteinuria are recognised phenotypic features associated with pathogenic *WT1* variants^36,37^. The recruiting clinician considered that the variant likely played a role in the patient’s renal disease and deemed it clinically reportable due to its potential relevance for both patient management and familial risk assessment.

In conclusion, our findings demonstrate that early parental mosaicism can be systematically detected from standard-depth trio WGS using careful bioinformatic filtering. This allowed us to survey PZM patterns at scale, across the largest number of individuals to date, and compare them to DNMs. Although rare, these events can cause rare conditions and have direct implications for recurrence risk estimates. As most remain undetected by current clinical pipelines, incorporating PZM detection into routine analysis could address a clear diagnostic gap and improve recurrence risk predictions for affected families.

## Data Availability

There are restrictions to the availability of the catalogue of postzygotic mutations described in this manuscript due to Genomics England policies on the export of individual-level information. This catalogue can be accessed from the Genomics England research environment upon request to join the Genomics England research network at https://www.genomicsengland.co.uk/join-us/academics. From the Genomics England research environment, our catalogue of postzygotic mutations and its associated code can be found in the following path: /re_gecip/shared_allGeCIPs/parental_pzm_cat.

## Declaration of interests

The authors declare that they have no competing interests.

## Acknowledgements

We thank the Wellcome Trust for funding this research. R.R. and H.C.M were supported by the Wellcome grant 220540/Z/20/A. This research was made possible through access to data in the National Genomic Research Library, which is managed by Genomics England Limited (a wholly owned company of the Department of Health and Social Care). The National Genomic Research Library holds data provided by patients and collected by the NHS as part of their care and data collected as part of their participation in research. The National Genomic Research Library is funded by the National Institute for Health Research and NHS England. We also thank Graciella Martin Rijo for generously sharing her views and suggestions on data visualization. For the purpose of open access, the authors have applied a CC-BY public copyright licence to any author accepted manuscript version arising from this submission.

## Author contributions

O.I.G.S. conducted the analyses and drafted the manuscript; R.S., M.T.I.B.; M.H.P. advised on QC or specific analyses and contributed to the generation of figures; J.A.S. provided clinical interpretation; A.S. provided scientific advice, H.C.M. and R.R. jointly supervised the study.

## Notes

### Competing Interest Statement

The authors have declared no competing interest.

### Author Declarations

This project relied on data collected and processed by the Genomics England 100,000 Genomes Project. Ethical approval for the 100,000 genomes project was given by the Health Research Authority of the National Research Ethics Service Committee East of England (REC: 14/EE/1112; IRAS: 166046). Access to this data was granted to OIGS through an application to the join Genomics England Clinical Interpretation Partnership (GeCIP) and the research registry project ID: 628.

