## Supplemental methods and figures for "Landscape of parental postzygotic mutations in >11,000 rare disease trios"

**Supplementary Methods**

**Section 1. Identification of candidate parental postzygotic mutations**

We accessed a comprehensive set of 14,446,487 unfiltered Mendelian inconsistencies, generated using the Platypus variant caller[15] from blood whole genome sequencing (WGS) data across 13,928 family trios, consisting of both parents and one offspring[1]. Variant calling was performed on a per trio basis, with each inconsistency representing a site where the offspring is heterozygous (HET) for the alternative (ALT) allele while both parents are homozygous for the reference (REF) allele. When multiple offspring were recruited for a given family, variant calling was performed separately for each offspring, meaning a given site could be flagged multiple times across siblings.

From this initial set, we retained only autosomal (chr1-22) single nucleotide variants (SNVs; A, T, G, C alleles) called on GRCh38-mapped genomes (n = 5,739,417). We excluded all trios previously identified to have biological or technical sources of germline hypermutation[2] yielding 5,720,018SNVs across 11,996 trios, involving 21,644 unique parents.

From these, we excluded 1,473,685 genotypes with zero ALT reads in both parents, reasoning that these are consistent with true *de novo* germline mutations transmitted to the child rather than parental PZMs, keeping 4,246,333 genotypes . For each trio, we excluded genotypes where the ALT allele had any coverage (>0 reads) in both parents (n = 1,628,565), retaining 2,627,735 genotypes that were considered candidate postzygotic mutations (cPZMs) for downstream filtering.

Additionally, since a single family could contribute multiple trios, we de-duplicated instances where the same variant was identified in the same parent across different offspring. After this step, we kept 2,574,260 cPZMs genotypes spanning 753,618 unique genomic positions, across 21,644 parents (mean = 118.93, Ts/Tv = 1.48).

**Section 2. Bioinformatic filtering of candidate parental postzygotic mutations**

We applied a rigorous, multi-step filtering pipeline to curate a high confidence set of candidate parental postzygotic mutations. Although 21,644 unique parents were originally considered, a total of 123 were removed due to missing individual or trio-level metadata. Hence, mean cPZMs per person were calculated based on a total of 21,521 parents.

1. **Removal of problematic genomic regions:** We excluded variants overlapping genomic regions prone to mapping errors, including interspersed repeats, low complexity regions, centromeres, segmental duplications, alternative haplotypes, and genome assembly patches. These regions were identified from the relevant GRCh38 tracks in the UCSC table browser[3]. This step reduced the dataset to 237,880 cPZM genotypes over 108,468 unique positions, across 21,456 parents (mean = 11.05, Ts/Tv = 0.86).

2. **Filtering common variants:** We removed variants with a global minor allele frequency (MAF) ≥ 1% in the 1000 Genomes Project (Phase 3)[4], resulting in 203,195 cPZM genotypes at 96,030 unique positions across 21,072 parents (mean = 9.44, Ts/Tv = 0.77).

3. **Excess coverage filter:** To remove potential sequencing artefacts, we excluded variants with site depth above an individual-specific upper threshold representing the top 1% of the expected coverage distribution, calculated using $qpois()$ in R ($lambda$ = mean whole-genome coverage; $lower.tail = FALSE$). We also removed 1,097 cPZMs from 117 parents with missing sequencing coverage metadata. After filtering 197,482 cPZM genotypes at 93,999 sites remained across 20,849 parents (mean = 9.17, Ts/Tv = 0.75).

4. **Minimum read coverage support filter:** Using $bamreadcount$[5], we calculated strand-specific ALT allele coverage for each cPZM and retained only variants with at least one ALT read on both strands. This yielded 77,548 cPZMs at 26,518 positions

across 14,898 parents (mean = 3.60; Ts/Tv = 0.69). Notably, 7,120 cPZMs had been assigned ALT = 1 in Platypus due to stringent filtering on windowed read support (see $bad - reads$ parameter[6]), which had led the GEL *de novo* pipeline to classify some of these mutations as candidate DNMs due to their apparent absence from parental blood.

5. **Variant allele fraction (VAF) consistency filter:** we used $binom.test()$ in R to assess whether parental VAFs were significantly <0.5 (one-sided test), and offspring VAFs consistent with 0.5 (two-sided test). We excluded variants failing either test (p >0.05 for parental VAFs, and p ≤0.05 for offspring VAFs), and 26 cPZMs from 6 parents with missing offspring read data. This retained 23,651 cPZMs at 12,325 unique positions across 8,848 parents (mean = 1.09, Ts/Tv = 0.77).

6. **Read quality based on edit distance filter:** We calculated the median edit distance (NM tag) of ALT supporting reads using $samtools\ view$ and $samjdk.jar$[7]. Reads with mapping quality <20 or matching the bitflag (--excl-flags) 3844 (i.e., unmapped reads, not primary alignments, reads failing vendor quality checks, optical and PCR duplicates, and supplementary alignments) were excluded. We kept variants with median NM ≤4, yielding 4,733 cPZMs at 2,796 sites across 3,955 individuals (mean = 0.21, Ts/Tv = 1.43). Details on the derivation of the NM threshold are described in **Supplementary Methods - Section 3**.

7. **Variant-specific error filter:** We estimated background error rate by pooling reads from 2,500 randomly selected unrelated parents (**Supplementary Methods - Section 4**). Variants with background ALT rate >0.00112 (matching the Illumina HiSeq 2500 error rate[8]) were excluded. This step retained 1,501 cPZMs at 1,492 sites across 1,401 parents (mean = 0.069, Ts/Tv = 1.68).

8. **Population frequency-based filtering:** We annotated the remaining cPZMs with their internal GEL cohort and gnomAD v3 population frequencies[9]. Only 9/1,501

variants were shared across multiple unrelated GEL parents. For external AFs, we found that 569/1,501 cPZMs (50.4%) were present in gnomAD. Only 98 of these variants (6.5%) were present at AFs >0.1%. To set an expectation of population allele frequency for *de novo* variation, we compared the gnomAD AFs from cPZMs with those from 778,347 filtered DNMs from the GEL cohort, previously reported by Kaplanis et al., 2022[24]. We observed that ~69% of DNMs were absent from gnomAD and 26% were present at AFs ≤0.1% (**Supplementary Figure 7**). However, we found significant differences between the proportion of variants absent from gnomAD and those present in gnomAD at AFs ≤0.1% between DNMs and cPZMs (single sample proportion test, p-value = $2.18 \times 10^{-09}$, and $2.18 \times 10^{-05}$ , respectively), indicating possible contamination of cPZMs with inherited HET variation.

To test for contamination by inherited HETs or sequencing errors, we compared VAFs for cPZMs across allele frequency bins. Variants absent from gnomAD or with AFs ≤0.1% had significantly lower VAFs than common variants (Wilcoxon rank-sum test, p = $2.2 \times 10^{-16}$ ; **Supplementary Figure 8A**). cPZMs shared by multiple parents in the dataset also had higher VAFs than cPZMs seen in just one parent (which we refer to as 'singletons') (Wilcoxon rank-sum test, p = $3.23 \times 10^{-07}$ ; **Supplementary Figure 8B**). Despite the ultra-rare group showing the lowest parental VAFs, the mean VAF of singleton variants absent from gnomAD was significantly lower than that of variants present at any AF in gnomAD (one way ANOVA and Tukey HSD post-hoc testing adjusted p-value ≤0.05; **Supplementary Figure 8C**). In contrast, among cPZMs shared across unrelated individuals in the GEL cohort, there was no difference in mean VAF between those absent from gnomAD versus present in any of the tested allele frequency bins in gnomAD (one way ANOVA and Tukey HSD post-hoc testing adjusted p-value > 0.05; **Supplementary Figure 8C**).

Based on the described observations, and to prevent the inclusion of putative HET variants, we first removed variants having a gnomAD AF >0.1% or shared among unrelated parents, retaining 1,389 cPZMs across 1,302 parents (mean = 0.064, Ts/Tv = 1.60).

9. **Empirical HET VAF threshold-based filter:** To remove putative heterozygous variants, we first estimated an individual-specific lower VAF bound for true HET sites using parental sequencing data. To do this, we started by obtaining germline genotypes for parents with at least one singleton cPZM (n = 1,302) from aggregated germline variant calls (AggV2) across 78,195 GEL participants in plink .pgen format, processed as described by Kousathanas et al. (2022)[40]. Genotypes were available for 1,293 individuals, as not all GEL participants were included in the aggregated dataset[41]. Using plink2, we generated an A-transposed genotype matrix (.traw) for SNPs with allele frequency between 0.2 and 0.8, identified sites called heterozygous (0/1) in at least one individual, and counted their occurrences among the 1,293 individuals. For each individual, we selected two SNPs per autosome (44 SNPs total), prioritising those most common in the subcohort. This yielded 74 unique SNPs, of which at least 44 were shared by ≥90% of individuals (**Supplementary Figure 9A**). We calculated VAF for each SNP using $bamreadcount$[5] (minimum base and mapping quality 20) and defined the lower bound for HET SNPs as 2.5 times the standard deviation of VAF across all SNPs in that individual (**Supplementary Figure 9B**). For nine individuals lacking SNP data, we used the mean threshold from the others (mean = 0.27).

   We then applied this threshold to the 1,389 remaining cPZMs, testing whether their VAFs fell significantly below the individual-specific lower bound using a one-sided $binom.test()$ (**Supplementary Figure 10A**). Variants with FDR-adjusted p-value >0.05 were excluded, leaving 1,015 high-confidence cPZMs (**Supplementary Figure 10B**) across 953 parents (mean VAF = 0.046, Ts/Tv = 1.57). No significant difference in parental VAFs was observed between candidates absent from versus present in gnomAD (Wilcoxon rank-sum test, p = 0.98).

A summary of the number of cPZMs per individual found across the whole cohort is shown in **Supplementary Figure 2**.

This final PZM catalogue had a Ts/Tv ratio of 1.57, which is lower than that expected from WGS variant calling (2-3[10]); however, our ratio is similar to that estimated for early PZMs reported by Sassani et al., 2019 (Ts/Tv = 1.6)[11]. Previous studies have shown that somatic mutation catalogues are expected to have a lower Ts/Tv ratios due to different cell selection processes operating in the relevant tissues[12]. Importantly, active and passive selection processes are known to occur prior to the differentiation of the epiblast[13], where PGCs originate[14].

**Section 3. Estimating an empirical maximum NM value for single nucleotide variants in GEL data**

The NM metric, which is produced during the alignment step of sequencing and can be extracted from the CIGAR string, represents the number of mismatches per read[15]. To calibrate a hard threshold for median NMs in cPZMs, we extracted these values from a set of high quality common polymorphic sites. Specifically, we extracted median NMs for a set of common SNPs overlapping with high-quality mapping regions of the human genome (GRCh38 reference) from parents carrying cPZMs. To do this, from the gnomAD v3.1.1 variant set[9] and for each autosomal chromosome (chr1-22), we extracted a random selection of 1000 bi-allelic SNPs (i.e. min and max alleles = 2), which we then intersected with high-confidence mapping coordinates produced by the Illumina's Platinum Genomes Project[16,17]. These genomic regions are known to produce consistent variant calls, as these have been validated through consistent inheritance patterns across a three-generation pedigree[16,17]. To retain only common SNPs likely to be shared by most individuals regardless of ancestry, we set a filter on gnomAD global allele frequency >=0.8.

As the purpose of calculating the median NM per variant is to approximate an expected number of mismatches in reads carrying the ALT allele in short read sequencing data, high quality SNPs obtained from previous steps were first compared against genotypes of the *de novo* cohort trio parents (n parents = 15,125). We checked that the alternate allele in the selected SNPs was found in heterozygous or homozygous form in at least 80% of the cPZM carriers. This was done using readily available aggregate VCF files containing filtered germline variant calls from 78,195 participants of the 100,000 genomes project[18], which includes all of the family trio parents studied in this section. Most of the selected SNPs were at least heterozygous in 80% of the family trio parents (n ~ 900 SNPs per chromosome). To reduce computing times, we only kept the 200 most common SNPs (i.e. most frequent in the cPZM carrier parents) per chromosome, and a random selection of 800 individuals for further testing. Between 4,397 and 4,600 (median = 4,600) genome-wide SNPs were tested per individual. In the same way to cPZM genotypes (see **Supplementary Methods - Section 2**), we extracted reads mapping to the ALT allele in each of the selected SNPs and calculated the median NM per site and individual. We found that >98% of high-quality SNPs had a median NM ≤4 (**Supplementary Figure 11**), so we used this as a hard threshold to filter cPZMs genotypes.

**Section 4. Generating variant-specific error rates for genomic sites harbouring cPZMs**

The variant-specific error rate is an empirical measure of the expected probability of observing a read with a given allele at a specific genomic position under random noise. In this context, it reflects either sequencing errors or common variation in the GEL cohort, with low 'error rate' variants being a product of the first instance, and variants with high 'error rate' being a product of the second.

To obtain the variant-specific error for a given genomic position, we pooled mapped reads across 2,500 randomly selected unrelated trio parents, and calculated the proportion of reads mapping to each single nucleotide allele (A, T, C, G, -). A list of unrelated individuals was readily available in the Genomics England data resources[19], and was produced by pairwise pruning of up to third-degree related individuals (e.g., cousins and great-grandparents) based on genetic relatedness scores obtained using the KING-robust algorithm. Random individuals were selected using the R $dplyr::sample\_n()$ function setting $replace = FALSE$. Reads were pooled using the R $deepSNV::loadAllData()$ function[20]. Base and mapping quality were set to a minimum of 20, and reads with the bitflag 3844 (i.e., unmapped reads, not primary alignments, reads failing vendor quality checks, optical and PCR duplicates, and supplementary alignments) were excluded. The variant-specific error rate distributions obtained using this method for the 4,733 cPZMs remaining after step 6 of the filtering are shown in **Supplementary Figure 12**.

### Section 5. Characterisation of PZMs

#### A. Comparing average PZMs per individual in this study versus the literature

For Rahbari et al., 2016, we consulted the "Table 1" found in their main text, and obtained the number of autosomal (chr1:22) single nucleotide PZMs assigned to each parent in the three families included in this study. PZM counts for each parent included any PZM entry where the ALT allele could be detected in siblings and in the parental blood (SM and SP), or in the parental blood alone (M or P). In total, this study reports 24 PZMs meeting the above criteria across six parents, corresponding to an average of 4 PZMs per parent.

For Sassani et al., 2019, we consulted their online supplementary material (https://github.com/quinlan-lab/ceph-dnm-manuscript), specifically the "gonosomal.dnms.summary.csv" table. This included all DNMs identified to have originated postzygotically across 69 individuals. To obtain the mean per individual, we summed the "snv_autosomal_dnms", which totalled 442 autosomal single nucleotide PZMs and corresponded to an average of 6.40 PZMs per individual.

For our study, we obtained the global average across all individuals that were not intentionally excluded (n = 123), totalling 21,521 individuals out of an initial selection of 12,644 parents. Individuals were excluded due to: a) presenting a form of true or false positive hypermutation, as reported by Kaplanis et al., (2022)[2], or b) participant ID inconsistencies in GEL data that prevented indexing metadata. Altogether, we identified an average PZM mutation rate of 0.08 PZMs per parent.

#### B. Association between parental ages at conception and trio DNMs and PZMs

DNMs detected in a trio are known to be associated with parental ages at conception, since this type of mutation accumulates with age in single gametes and gamete progenitor cells in the case of males[21]. Since parental PZMs are expected to have originated during the parental embryonic development, a correlation between the total number of parental PZMs detected in an offspring and the parental age at conception is not expected. To corroborate this in our data, for each trio we obtained PZMs per trio, by summing the PZMs ascertained from the mother and father. We accounted for the de-duplication of PZMs performed earlier in our filtering pipeline, and we assigned sibling-shared PZMs to the corresponding offspring (n = 20). For all offspring carrying at least one PZMs (n = 955), we obtained the unphased DNMs counts. These corresponded to a set of pre-filtered and bioinformatically ascertained per trio DNMs, previously reported by Kaplanis et al. (2022). We then used a generalised linear regression model from the “Poisson” family to model mutation counts (either total DNMs or PZMs) as a function of parental ages at conception while controlling technical covariates that could have affected the mutation calling process, as follows:

$$mutation\ counts\ = \beta_0\ +\ maternal\ age\ at\ conception\ \bullet\ \beta_1\ +$$

$$+\ paternal\ age\ at\ conception\ \bullet\ \beta_2\ +$$

$$mean\ sequencing\ depth_{\ mother}\ \bullet\ \beta_3\ +mean\ sequencing\ depth_{\ father}\ \bullet\ \beta_4\ +$$

$$mean\ sequencing\ depth_{\ offspring}\ \bullet\ \beta_6\ +$$

$$percent\ aligned\ reads_{\ mother}\ \bullet\ \beta_5\ +percent\ aligned\ reads_{\ father}\ \bullet\ \beta_6\ +$$

$$percent\ aligned\ reads_{\ offspring}\ \bullet\ \beta_7\ +\ \varepsilon$$

We found no significant association between PZM counts per trio and maternal or paternal ages at conception (p-value = 0.54, p-value = 0.420, respectively), while these were significantly associated with DNM counts (maternal effect = $7.2 \times 10^{-3}$, p-value = $6.37 \times 10^{-14}$; paternal age effect $1.8 \times 10^{-2}$, p-value < $2 \times 10^{-16}$).

#### C. Testing genomic clustering of PZMs and DNMs

To assess whether PZMs (or DNMs) are clustered in specific genomic regions, we performed a permutation test on binned mutation counts. We first divided the genome into consecutive two-megabase (Mb) windows and counted the number of mutations in each bin. To generate the null distribution, we randomly reassigned the total number of observed mutations across all bins, while accounting for differences in bin size due to mappability and filtering.

Specifically, we adjusted for the callable size of each bin by subtracting the number of bases overlapping regions excluded during filtering (e.g., segmental duplications, centromeres). We then computed a per-bin probability of receiving a mutation as the callable size of the bin divided by the total callable genome size. This weighted random assignment was repeated 10,000 times to generate a null distribution of mutation counts per bin. Finally, we computed one-tailed empirical p-values per bin as the proportion of permutations in which the number of mutations assigned to a bin was equal to or greater than the observed count. We corrected our p-values to account for multiple testing using the “Benjamini-Hochberg” method.

#### D. Testing the association of genomic features with clustered PZMs and DNMs

Using the bin definition outlined in the previous section, we tested whether mutation-enriched bins had differential genomic features as defined by GC content and replication timing. We annotated GC content per bin by extracting the GRCh38 sequences from each bin using the R libraries bin GenomicRanges and Biostrings[22,23]. For each bin, we counted the total number of non “N” nucleotides and determined the GC content as the proportion of “G” and “C” nucleotides out of this total. Replication timings were obtained from Massey et al., (2019)[24]. These were derived from the GM12878 lymphoblastoid cell line from which replication timing was inferred for the S-phase sorted cells, using G1-sorted cells as baseline[24]. This replication timing profile was selected on the basis of being representative of at least 50% of the replication timings across cell types[25,26].

We tested whether GC content, replication timing, and callable size per bin predicted the likelihood of a bin being significantly enriched for PZMs or DNMs using a generalized linear model with a binomial link function. Coefficients from this regression represent the change in odds of bin enrichment significance per unit increase in each predictor variable. Bin enrichment for each mutation type was determined as follows. For PZMs, we define as significant those bins having a nominal permutation p-value ≤ 0.05 (n = 49). For DNMs, we define as significant those bins having an FDR-adjusted permutation p-value <= 0.05 (n = 236).

#### E. Mutation spectra annotation of PZMs

We annotated each SNV using according to the pyrimidine change caused by the ALT allele (i.e., C>A, C>T, C>G, T>A, T>C, T>G). In addition, we differentiate between C>T sites occurring in CpG sites (CpG>TpG) and elsewhere in the genome, to account for the differential number of transitions of these genomic contexts[27,28]. Using the seven pyrimidine substitution classifications outlined above, we calculated proportion differences across sexes and DNM events. For each group comparison (paternal vs maternal PZMs; DNMs vs PZMs), we estimated if proportions of mutations assigned to each mutation category were significantly different using a two-sample proportion test. P-values for each experiment were adjusted to account for multiple comparisons using the FDR method.

To test if PZMs and DNMs have a different mutational origin, as defined by their mutation spectra, we performed mutational signature extraction using SigProfiler[29]. For this, we annotated the pyrimidine substitutions obtained above with their 5’ and 3’ contexts and generated counts matrices for the resulting combinations (n = 96). We used this as input for SigProfiler extractor to estimate *de novo* mutational signatures. We deconvoluted the resulting signatures into reference signatures reported in the COSMIC[30] database using the relevant method in the SigProfiler software.

#### F. Consequence prediction, functional annotation, and identification of potentially clinically relevant PZMs

We used the Ensembl variant effect predictor (VEP, version 112) to generate functional effect predictions for PZMs[31]. We used bcftools +split-vep plugin[32] to keep the transcript for which the worst consequence was found. VAF differences across VEP categories were assessed using a linear regression model of the gamma family to account for the skewed VAF distribution of PZMs. We first modelled VAF as a function of six VEP categories, namely: LoF, missense, splice, synonymous, UTR, and non-coding variants. We also modelled VAF as a function of variants found overlapping coding exons or not (exonic, non-exonic variants). None of the tested categories was significantly associated with PZMs VAF (p-value >0.05).

We also tested enrichment of PZMs across genomic regions predicted to have specific functions. For this, we assigned genomic coordinates (GRCh38) to one of three categories: transcriptional units (i.e., introns, exons, UTR regions), putative regulatory regions (5000 bases upstream transcription start sites or downstream transcription termination sites), and intergenic regions. Coordinates for these elements were inferred from exon, intron, and UTR coordinates from the Refseq MANE select coding transcripts, obtained from the UCSC table browser[33]. Then we used the genomic association tester (GAT) algorithm to test if the observed number PZMs overlapping each annotation was significantly different from expectation under random chance[34].

To identify PZMs with potential clinical value, we looked at those whose VEP functional impact was classed as either HIGH or MODERATE (n = 21), and extracted the HUGO symbols for the affected genes. Then, we intersected these genes with PanelApp genes (n = 6,461)[35], keeping 12 genes (one per PZM variant) for further inspection. We then kept PZMs in genes found in gene panels in the "green" category (i.e., those with definitive evidence of causality for disease[35]), and that have a dominant effect in disease (i.e., monoallelic inheritance) according to the same resource. This resulted in the identification of five genes *(VAMP2, WT1, DYNC1H1, PHIP,* and *NLRP3*). From these, we followed up genes whose variant carrier was classed as "undiagnosed" in GEL records. We discarded *NLRP3* since the proband was reported to have been diagnosed on the basis of a different gene variant. For the rest, we inspected individual phenotype records, and compared these against the phenotypes reported by PanelApp and in the literature. We discarded *PHIP* from further description since the patient's phenotypes were not a good match with Chung-Jansen Syndrome, associated with *PHIP* mutations[36].

18. Aggregated Variant Calls (AggV2) - Genomics England Research Environment User Guide https://re-docs.genomicsengland.co.uk/aggv2/.

19. AggV2 Principal Components and genetically inferred relatedness - Genomics England Research Environment User Guide https://re-docs.genomicsengland.co.uk/principal_components/.

20. Gerstung, M., Beisel, C., Rechsteiner, M., Wild, P., Schraml, P., Moch, H., and

Beerenwinkel, N. (2012). Reliable detection of subclonal single-nucleotide variants in tumour cell populations. Nat. Commun. *3*, 811. https://doi.org/10.1038/ncomms1814.

21. Moore, L., Cagan, A., Coorens, T.H.H., Neville, M.D.C., Sanghvi, R., Sanders, M.A., Oliver, T.R.W., Leongamornlert, D., Ellis, P., Noorani, A., et al. (2021). The mutational landscape of human somatic and germline cells. Nature *597*, 381–386. https://doi.org/10.1038/s41586-021-03822-7.

22. Lawrence, M., Huber, W., Pagès, H., Aboyoun, P., Carlson, M., Gentleman, R., Morgan, M.T., and Carey, V.J. (2013). Software for computing and annotating genomic ranges. PLoS Comput. Biol. *9*, e1003118. https://doi.org/10.1371/journal.pcbi.1003118.

23. Biostrings Bioconductor. https://bioconductor.org/packages/release/bioc/html/Biostrings.html.

https://doi.org/10.1016/j.xgen.2022.100179.

### Supplementary Figures

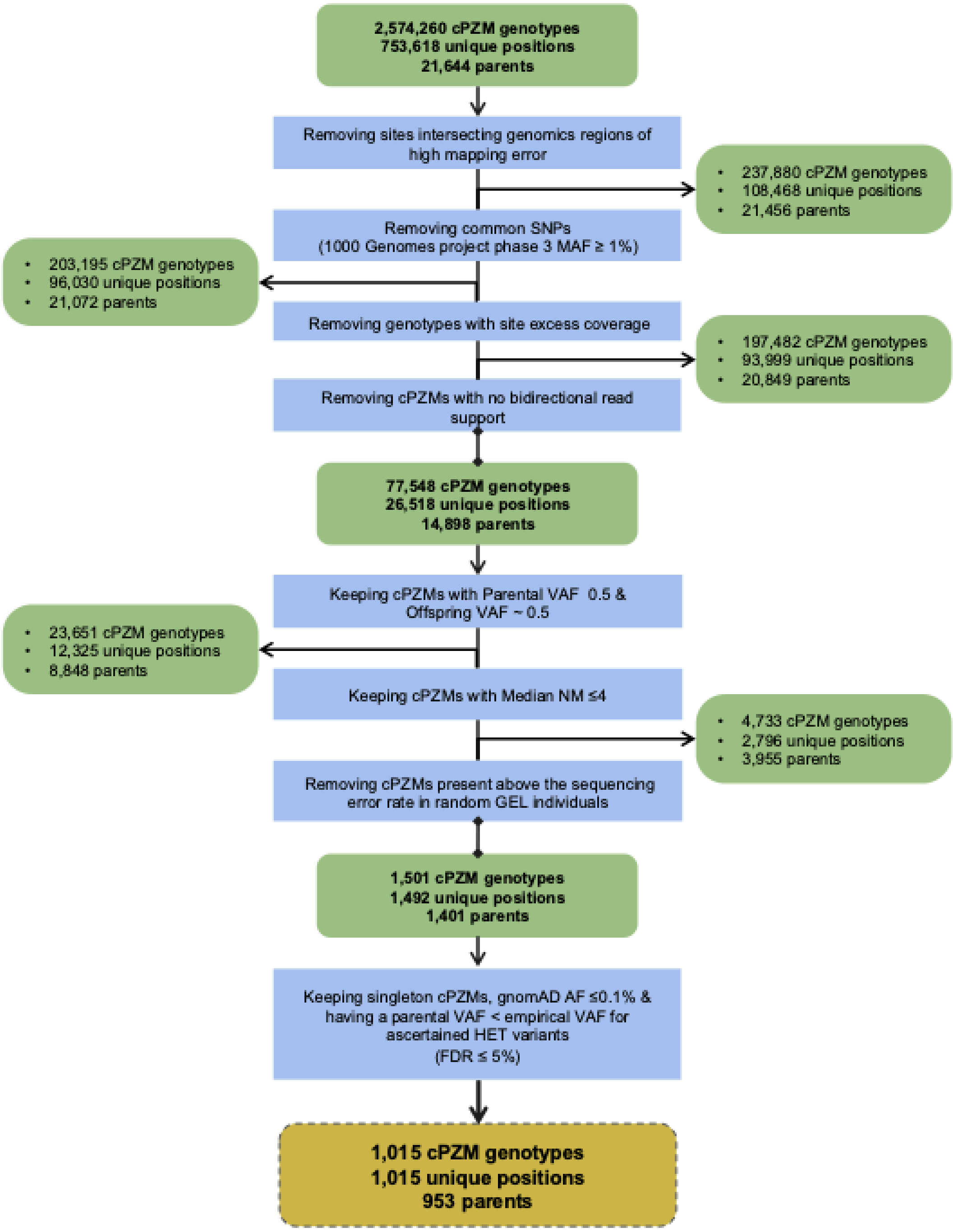


**Supplementary Figure 1. Summary of filtering steps for bioinformatic ascertainment of parental postzygotic mutation in the GEL family trio cohort.**

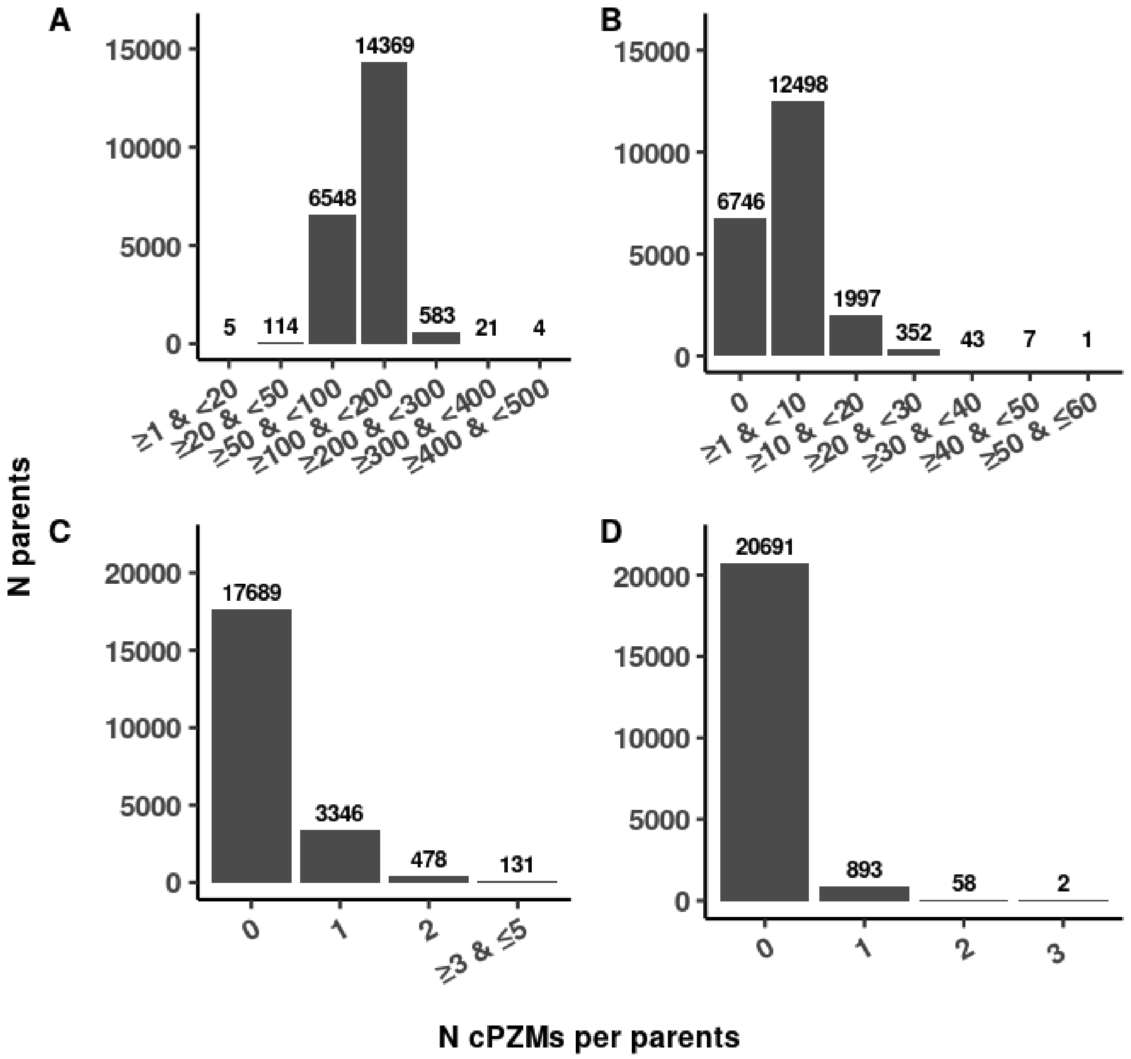


**Supplementary Figure 2. Number of cPZMs per parent across main filtering steps.**

Number of candidate cPZMs per individual across the whole initial cohort of 21,644 parents for: **A.** Initial selection of 2,574,260 cPZMs. **B.** 77,548 cPZMs remaining after removing variants intersecting genomic regions of high mapping error, reported in the 1kGP phase 3 (MAF ≥1%)[4], with a total site sequencing above individual sequencing-excess thresholds (as defined in filtering step 2 - **"**Excess coverage filter"), or supported by less than one read on each strand **C.** 1,501 cPZMs remaining after removing likely HET variants based on parental and offspring VAFs, having a median NM >4, or being above the median Illumina 2500 error rate in random GEL participants. **D.** 1,015 cPZMs remaining after removing variants with gnomAD AF >0.1%, shared across unrelated parents, or not being significantly different from the empirical lower-bound VAF threshold for ascertained HET variants (FDR >5%). Top numbers indicate counts per bin and variant type.

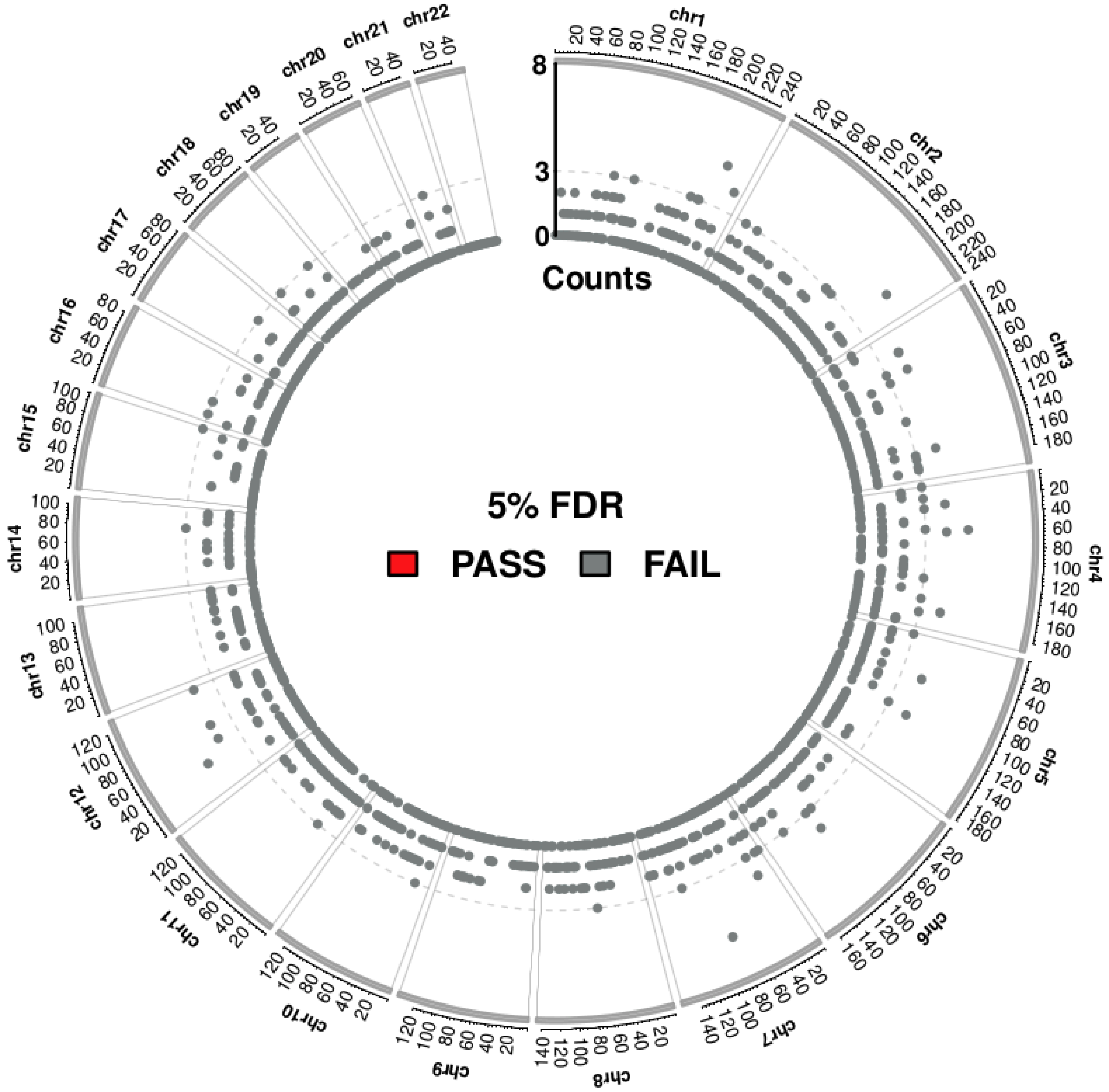


**Supplementary Figure 3. Genomic distribution of PZMs**

Each point represents cohort-wide PZM counts for a given non-overlapping 2Mb window. No bin was found significantly enriched after multiple testing significance correction.

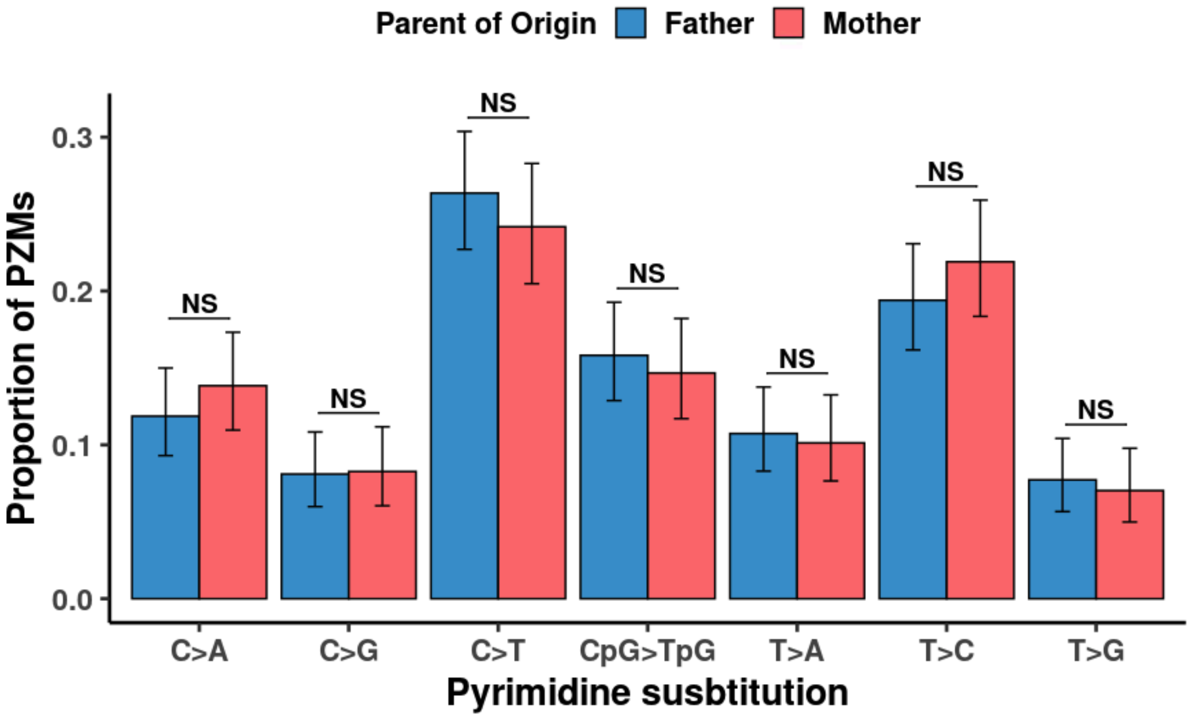


**Supplementary Figure 4. Spectral characteristics of PZMs stratified by sex.**

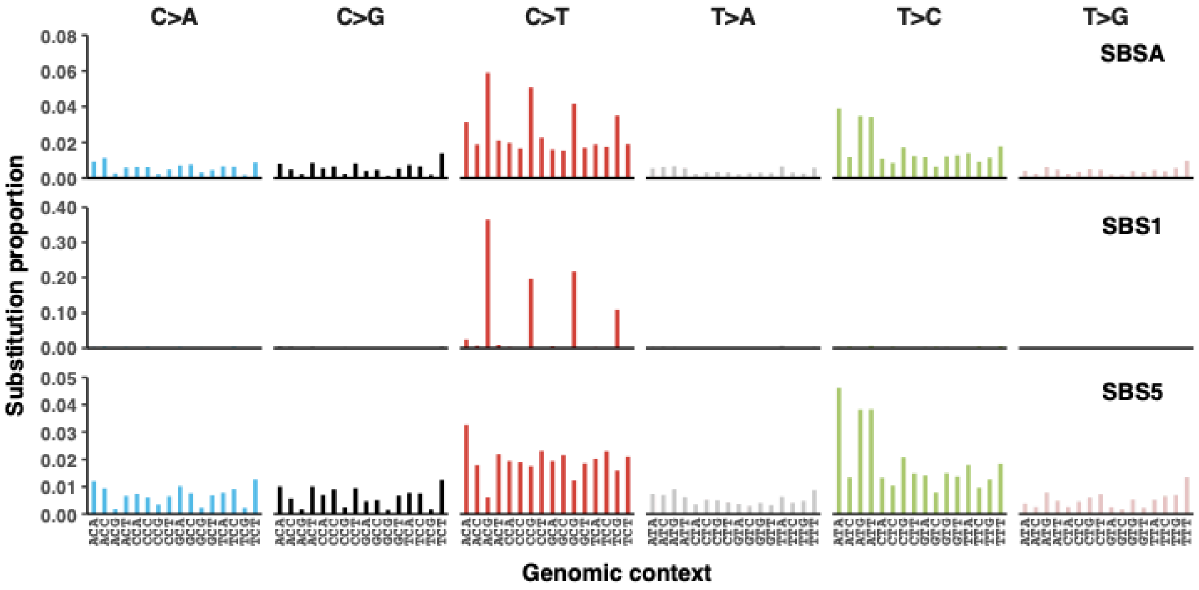


**Supplementary Figure 5. Mutational signature extraction and deconvolution of PZMs.**

Showing mutation proportions for *de novo* extracted signature profile from GEL DNMs and PZMs (i.e., SBSA), and SBSA deconvoluted into COSMIC signatures SBS1 and SBS5 (cosine similarity = 0.98).

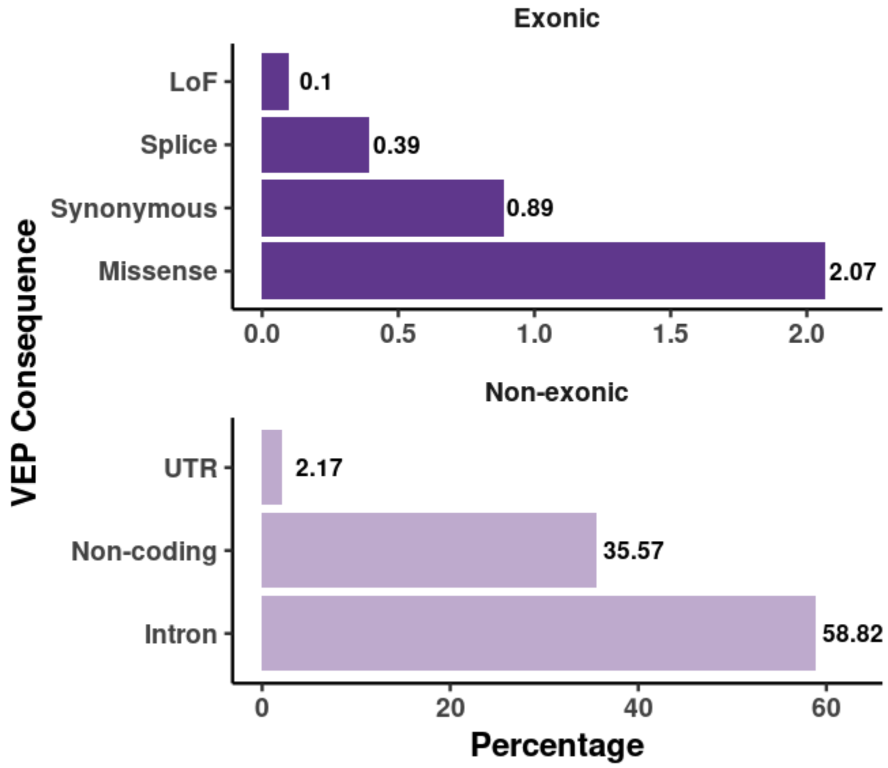


**Supplementary Figure 6. Consequence effect prediction for final cPZMs set (n = 1,015).**

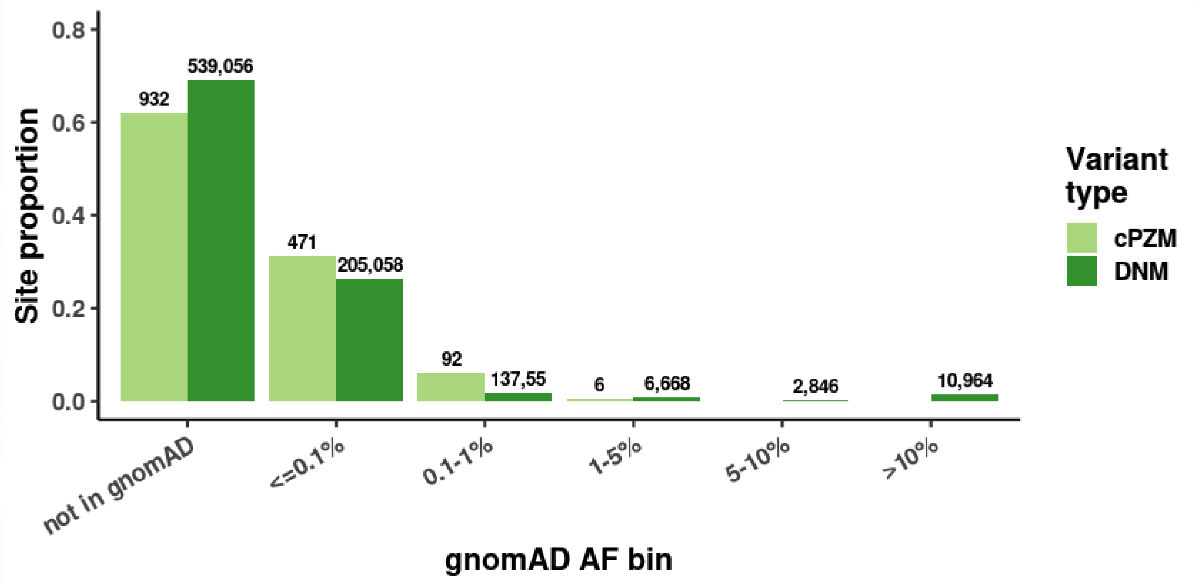

**Supplementary Figure 7. Distribution of population allele frequencies (gnomAD genomes v3).**

Showing binned gnomAD allele frequency distributions for 1,501 cPZMs remaining after step 1-7 filtering and filtered GEL DNMs per trio (stringent filter) reported by Kaplanis et al. [2]. Top numbers indicate counts per bin and variant type.

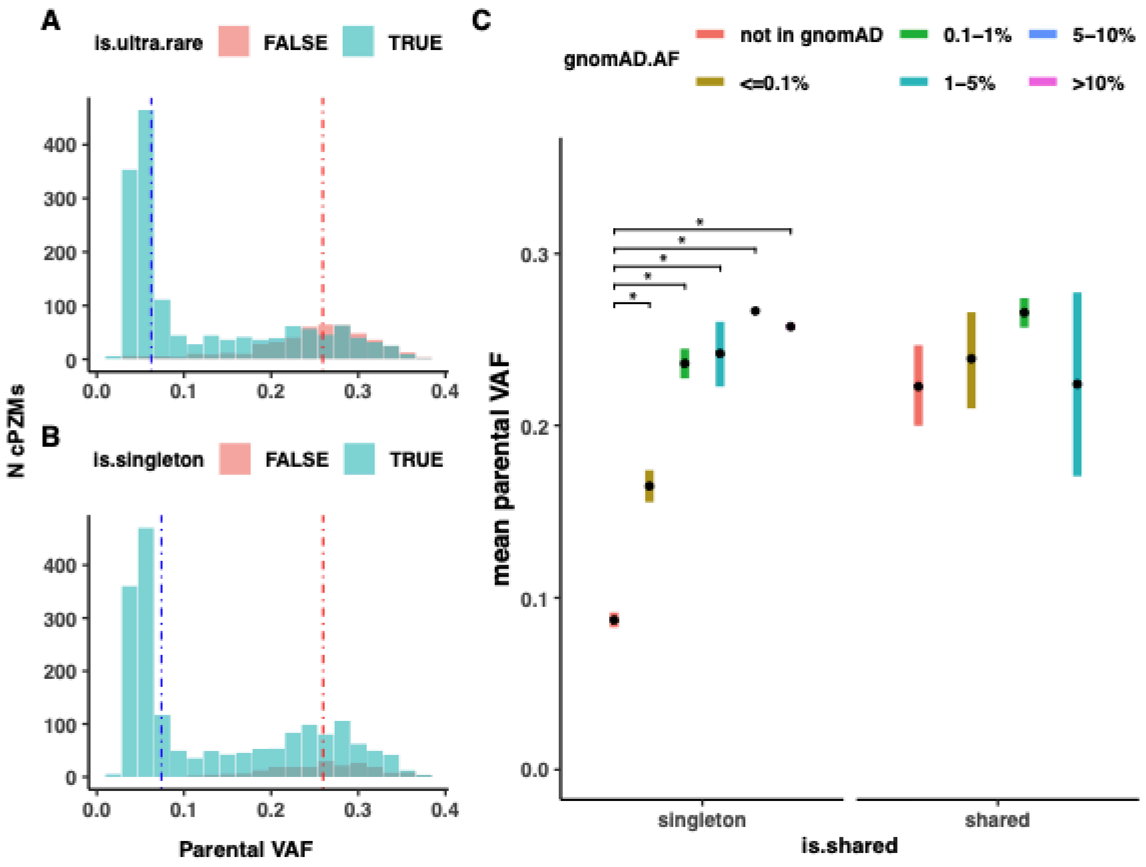


**Supplementary Figure 8. Parental VAF comparisons across population allele frequency bins for cPZMs passing filtering steps 1-7.**

**A.** Parental VAF of ultra-rare and non-ultra-rare cPZMs classes (n = 1,501). The ultra-rare group represents a composite between cPZMs absent from gnomAD or having a gnomAD AF ≤0.1%. **B.** Parental VAF of cPZMs found as singletons (i.e., found in a single parent) or shared across the 1,401 unrelated parents having at least one cPZM passing filtering steps 1-7. **C.** Mean parental VAFs for cPZMs stratified into six population allele frequency bins (gnomAD) and according to their presence across unrelated parents (i.e. singleton = seen in only one parent, shared = seen in one or more parent). Bars correspond to 95% confidence intervals for the mean estimate for each group. Horizontal lines and asterisks indicate significant VAF differences between groups (ANOVA and Tukey HSD post-hoc test adjusted p-values ≤ 0.05).

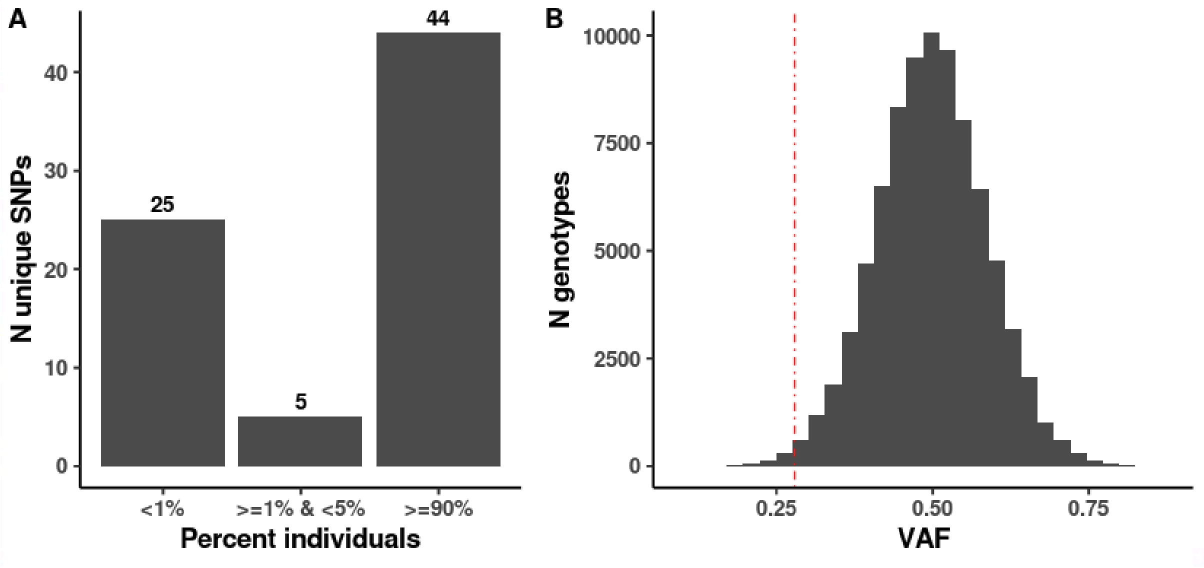


**Supplementary Figure 9. Description of SNPs selected to derive an empirical lower variant allele fraction (VAF) bound threshold for heterozygous (HET) variants.**

**A.** Number of HET SNPs shared across different proportions of the subcohort of interest (n = 1,293)**.** Top numbers indicate the number of individuals in each bar. **B.** VAF distribution for HET genotypes at the selected SNPs. The red line overlaps the mean lower-bound VAF threshold (mean VAF threshold = 0.27) across 1,293 individuals.

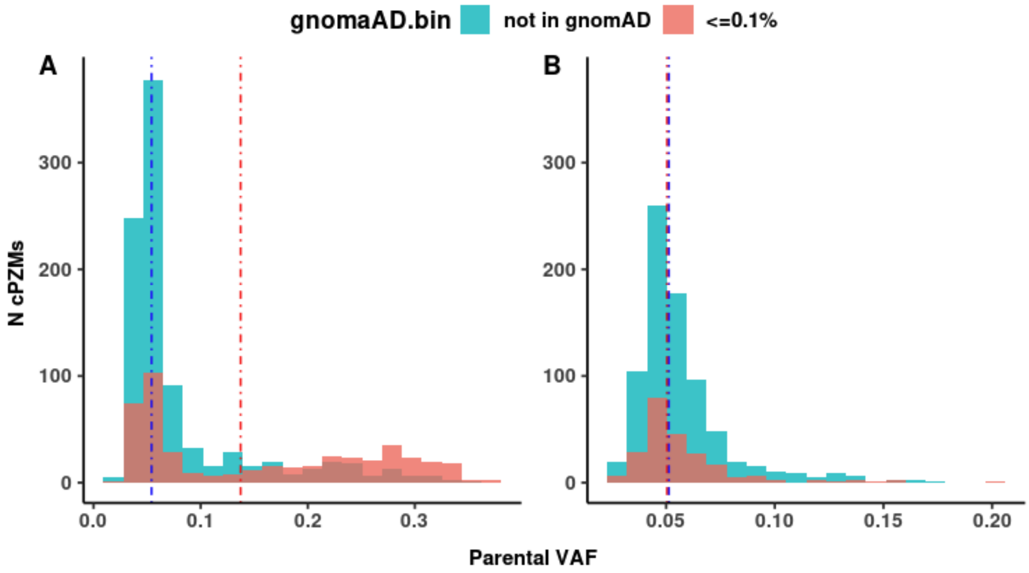


**Supplementary Figure 10. Parental VAF distribution comparisons for singleton cPZMs stratified by gnomAD allele frequency.**

**A.** All singleton cPZMs passing filtering steps 1-8 (n = 1,389). **B.** Singleton cPZMs after removing putative HET sites (filtering step 9; n = 1,015). Dotted lines intersect with median parental VAFs for cPZMs not found in gnomAD (blue), and found in gnomAD (red) in each panel. Median parental VAF is significantly different between groups before removing putative HETs (Wilcoxon rank-sum test p-value = $2.2 \times 10^{-16}$), and non-significant after filtering these sites (Wilcoxon rank-sum test p-value = 0.98).

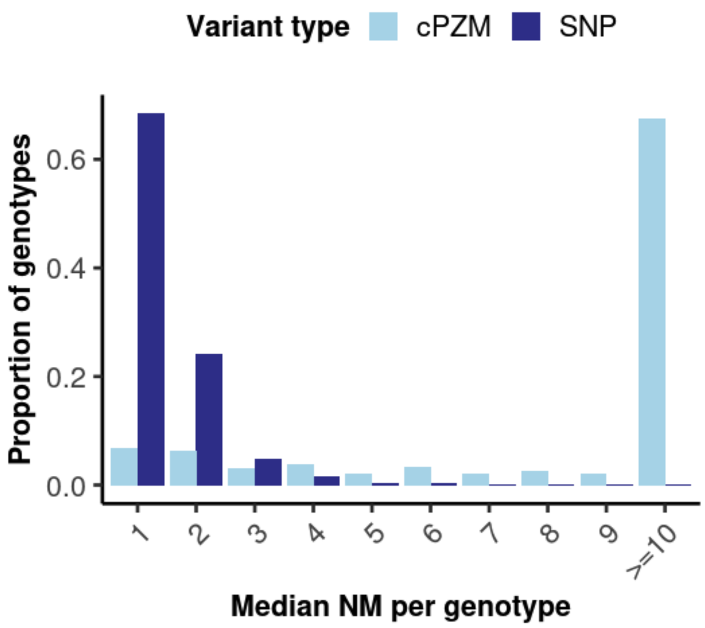


**Supplementary Figure 11. Distribution of median number of mismatches (NM) for reads mapping to cPZMs and GIAB SNPs.**

Showing proportions out of a total of 23,651 cPZMs passing filtering steps 1-5, and up to 4600 high quality SNPs.

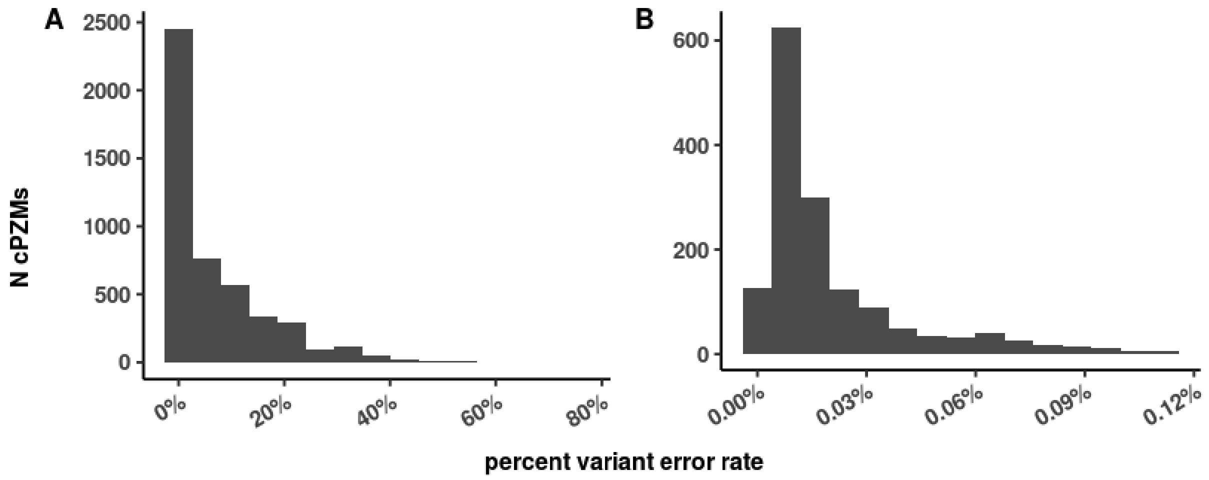

**Supplementary Figure 12. Variant-specific error rate distributions for cPZMs passing filtering steps 1-6.**

Showing variant specific error rates for 4,734 cPZMs **A.** passing filtering steps 1-6 **B.** passing filtering step 8 (n = 1,501). Error rate filtering threshold corresponds to the median Illumina HiSeq 2500 error rate (0.112%).

**Supplementary Tables**

| Embryonic cell division | Number of Cells | Expected cell proportion carrying the mutant allele | Expected variant allele fraction |
|---|---|---|---|
| 1 | 2 | 0.5 | 0.25 |
| 2 | 4 | 0.25 | 0.125 |
| 3 | 8 | 0.125 | 0.0625 |
| 4 | 16 | 0.0625 | 0.03125 |
| 5 | 32 | 0.03125 | 0.015625 |
| 6 | 64 | 0.015625 | 0.0078125 |
| 7 | 128 | 0.0078125 | 0.00390625 |
| 8 | 256 | 0.00390625 | 0.001953125 |
| 9 | 512 | 0.001953125 | 0.0009765625 |
| 10 | 1024 | 0.0009765625 | 0.00048828125 |

**Supplementary Table 1. Expected PZM VAFs and mutant cell proportions in adult tissues**

Simplified model assuming synchronous and equal cell division, a stable diploid genome, no selection on mutant cells, and an even distribution of mutant cells across adult tissues for PZM events occurring between the 1st and 10th cell division.
